# Severe COVID-19 pneumonia and barotrauma: From the frying pan into the fire

**DOI:** 10.1101/2021.02.12.21251479

**Authors:** Hariprasad Kalpakam, Sameer Bansal, Nithya Suresh, Samson Kade, Anmol Thorbole, Ravindra M Mehta

**Affiliations:** Department of Pulmonary, Critical Care and Sleep Medicine, Apollo Specialty Hospitals, Bangalore, India

**Keywords:** COVID-19, Lung Diseases, ARDS, Barotrauma, C-ARDS

## Abstract

**Aim:** COVID-19 pneumonia with ARDS (C-ARDS) has a high mortality. Preliminary reports indicate a higher incidence of barotrauma in patients with C-ARDS[1] both on invasive mechanical ventilation (iMV) and non-invasive ventilation (NIV) This study examines the incidence and risk factors for barotrauma and change in outcomes after barotrauma in patients with severe C-ARDS on positive pressure respiratory support (PPRS).

**Methods and materials:** This is a retrospective study of C-ARDS associated barotrauma over 5 months in patients on PPRS in a tertiary COVID care center. The type of barotrauma, intervention, related factors, such as type of respiratory support (iMV vs NIV), airway pressure prior to the occurrence of barotrauma, and post-barotrauma outcomes were analyzed.

**Results:** A total of 38/410 (9.3%) C-ARDS patients on PPRS [mean age 57.82 ± 13.3 years, 32 males (84.2%)] developed barotrauma. Of these, 20 patients (52.6%) were on NIV and 18 (47.4%) patients were iMV on standard recommended settings. The median P/F ratio of patients on MV at the time of barotrauma was 116.4 (IQR 72.4, 193.25). The details of barotrauma were as follows: 24 patients had pneumothorax (PTX), 2 had pneumo-mediastinum and 12 had subcutaneous emphysema. Overall, 24/38 (63.2%) patients, including 15/18 (83.3%) on MV succumbed to their illness. The barotrauma happened a median of 6.5 days (IQR 4.75,13) after admission and 15 days (IQR 10.25,18.0) from symptom onset. The median duration from barotrauma to death was 7 days (IQR 2.25, 8.0) and barotrauma to discharge (for survivors) was 12.5 days (IQR 8.0, 21.25). All patients received steroids and 11/38 (28.9%) received additional immunosuppression with tocilizumab.

**Conclusion:** A high incidence of barotrauma was seen in this large series of severe C-ARDS patients on PPRS. Barotrauma led to further deterioration in the clinical status leading to a fatal outcome in the majority of the MV patients, despite prompt treatment.

## Main paper

### Introduction

COVID-19 ARDS (C-ARDS) in the absence of effective therapy is associated with a high mortality, especially in patients with multiple co-morbidities and in the elderly. Positive pressure ventilation with PEEP has been used in ARDS management since the first description of ARDS in 1967 by Ashbaugh et al.[2] Barotrauma is a known complication of ARDS, prevented by standard ventilation strategies such as low tidal volume ventilation, appropriate PEEP and limiting plateau pressures ≤ 30 cm H_2_O. The incidence of barotrauma in mechanically ventilated (MV) patients is about 2.9 %. Chronic obstructive pulmonary disease (COPD), interstitial lung disease (ILD) and *Pneumocystis jeroveci* pneumonia (PJP) are associated with a higher incidence of barotrauma. Anzueto et al. reported an incidence of barotrauma of 10.0% in MV patients with chronic ILD.[3] Viral pneumonia caused by H1N1, SARS-COV1 and MERS-CoV with severe ARDS are also associated with barotrauma. A high incidence of barotrauma (17%) was reported in a case series of critically ill MERS-CoV patients.[4] Barotrauma is also associated with adverse outcomes in these patients and often portends a rapid downhill course.[3] In several observational studies on C-ARDS, a high incidence of barotrauma was reported. McGuinness et al reported a 15% incidence of barotrauma in 601 patients with COVID-19 requiring MV support. [5] Younger age and longer hospitalization were found to be risk factors for developing barotrauma. The high incidence of barotrauma in viral pneumonia especially with coronaviruses suggests a unique pathophysiology and clinical challenge different from bacterial pneumonia. Unlike other ARDS, C-ARDS barotrauma has been seen not only in MV patients, but across all categories of positive pressure respiratory support (PPRS). In this study, we report the incidence of, risk factors for, and outcome after barotrauma in C-ARDS patients on PPRS.

## Materials and methods

ICU patients with confirmed critical COVID-19 ARDS (C-ARDS) requiring respiratory support in the form of high flow nasal canula (HFNC) /non-invasive ventilator (NIV) or invasive MV (iMV) were included in the study. Barotrauma was defined as the presence of new onset extra-alveolar air, namely pneumothorax (PTX), pneumo-mediastinum or subcutaneous emphysema. [6] Barotrauma was discovered on a routine chest radiograph (CXR) done daily for patients on respiratory support or on urgent imaging (CXR, point-of-care ultrasound) done in the event of clinical deterioration. [Fig 2 (a) & (b)] The respiratory and ventilator settings (peak airway pressure, plateau pressure, PEEP and P/F ratio (PaO2/FiO2 ratio) preceding the barotrauma were noted. D-dimer levels was extracted from the medical records. Strategies for management of barotrauma included either observation or intercostal drainage (ICD), modification in ventilator settings to reduce mean airway pressures, tapering or discontinuing steroids and aggressive treatment of secondary infections. NIV support was replaced with HFNC or mask oxygen when possible depending on the patient’s clinical status. The main outcomes were analyzed as death or discharge from the ICU.

**Fig (1).**
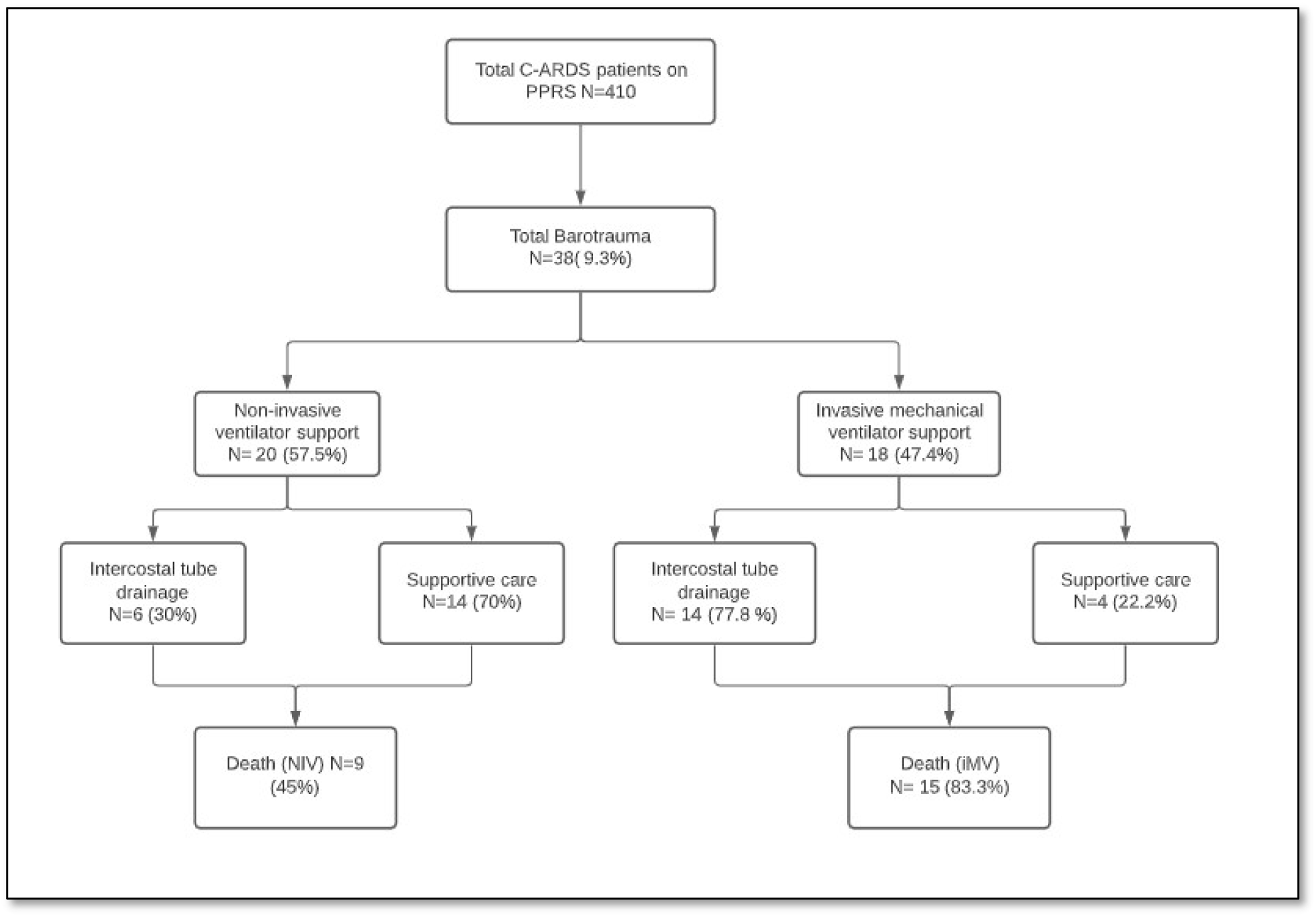
Flow-chart of patients on PPRS who developed barotrauma and eventual outcomes.

**Fig 2.**
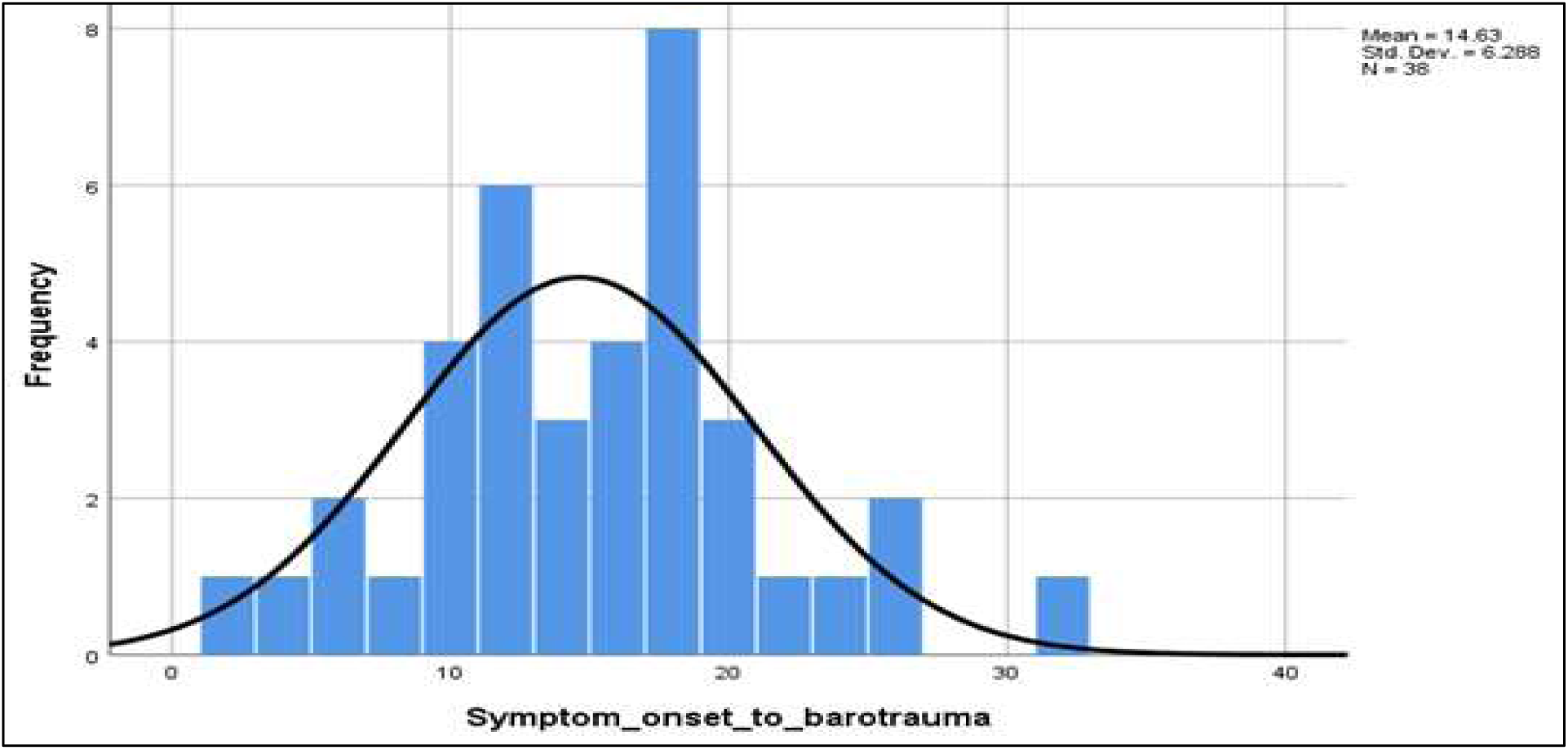
Symptom onset to barotrauma (Peak barotrauma events occurred between 10 & 20 days from the onset of symptoms)

### Statistical analysis

Descriptive statistics were followed; the continuous variables were expressed as mean ± SD or median ± IQR (Interquartile range) and discrete variables as rates and percentages.

## Results

There were a total of 38/410 (9.3 %) C-ARDS ICU patients on PPRS [mean age 57.89 ± 13.12 years, including 32 males (84.2%) who developed barotrauma. 20 (57.5%) patients were on NIV and 18(47.4%) patients were on iMV (Fig 1). The incidence of barotrauma in patients on positive pressure respiratory support (PPRS) was 11 %; (18/163) in the iMV group and 8.1 % (20/ 247) in the NIV group. Median settings preceding the barotrauma in ventilated patients were peak inspiratory pressure (P-peak) 33.5 cm H_2_O (IQR: 30,35.25), plateau pressure (P-plateau) 30.0 cm H_2_O (IQR 25.5, 31), positive end-expiratory pressure (PEEP) 8.0 cm H_2_O (IQR 6.0, 10), and tidal volume (V_T_) 420 ml (IQR 380, 420). The median P/F ratio of patients on iMV at the time of barotrauma was 116.4 (IQR 172.4, 193.25).

7 patients on MV at the time of barotrauma were on prone ventilation. The nature of barotrauma was as follows: 24 patients had PTX, 2 patients had pneumo-mediastinum and 12 patients had subcutaneous emphysema. The median D-dimer levels in all the barotrauma patients was 1159 (IQR 438, 2268.50), significantly higher in patients on iMV −1381.5 (IQR 914.5,2220). Interventions for barotrauma included ICD in 20 patients (52.6%) and supportive care for the rest. Imaging post-ICD tube placement revealed resolution/reduction of PTX in all but 2 patients. Overall, 24/38 (63.2%) patients including 15/18 (83.3%) on iMV succumbed to their illness despite appropriate treatment measures for the barotrauma. The barotrauma happened a median of 6.5 days (IQR 4.75,13) from admission and 15 days (IQR 10.25,18.0) from symptom onset. The median duration from onset of barotrauma to death was 7 days (IQR 2.25, 8.0) and barotrauma to discharge (for survivors) was 12.5 days (IQR 8.0, 21.25). Two patients had a prolonged air leak persisting more than a month.

## Discussion

In this observational study of severe C-ARDS patients on PPRS there was a high incidence of barotrauma (9.3 %), of different types. The rates of barotrauma are significantly higher than those reported in prior reports of ARDS of other etiologies but similar to other viral ARDS.[7] The barotrauma was seen in both NIV and iMV, though patients on iMV had a higher incidence (11% in iMV vs. 8.1% in the NIV group). The barotrauma occurred despite the use of standard ARDS ventilatory strategies (6-8ml/Kg tidal volume and plateau pressure ≤ 30 cm H2O) in the iMV group and similar TV targets in the NIV group. The incidence of barotrauma varies depending on the etiology of ARDS. Both SARS CoV1 and MERS-CoV were associated with a high incidence of barotrauma in patients with ARDS [4, 8], including patients on NIV. The SARS-CoV1 epidemic in 2002 revealed a barotrauma incidence of 6.6% −15% including patients receiving NIV.[9] In other non-infectious conditions, the incidence of barotrauma with iMV varies depending on the underlying lung disease, from 2.9% overall to 10% in ILD with respiratory failure. Ventilated ILD’s have a high incidence of barotrauma likely similar to the later stages of C-ARDS when fibrosis develops. The pathophysiology of C-ARDS barotrauma is multifactorial, and includes inherent disease aspects, and differing ventilation strategies, and other factors. Accordingly, the possible causes of barotrauma in C-ARDS can be divided into extrinsic and intrinsic factors (Table 1). Extrinsic factors as commonly understood include high inflation pressures, PEEP, and prolonged ventilation. The effect of high airway pressures on barotrauma is well established, although the precise levels (P-Peak & P-plateau) of airway pressure that led to barotrauma are less well defined.

**Table (1).** Intrinsic & extrinsic causes for barotrauma

| Intrinsic | Extrinsic |
| --- | --- |
| Severe ARDS | High inflation pressures |
| Time from Disease onset | PEEP |
| Necrotizing infections | Prolonged ventilation |
| D-dimer | Prolonged NIV and delayed initiation of MV |
|  | Medications (viz. steroids and other immunosuppressive medications) |

The relationship of ventilation parameters and incidence of barotrauma is not well-defined. Weg et al. noted higher values for PEEP and respiratory rate in 725 non-COVID ARDS (nCov-ARDS), compared to standard values as predictive factors for pneumothorax.[10] Conversely, Gammon et al. found no association between airway pressures and barotrauma in a similar population of nCov-ARDS. [11] Eisner et al using data from the ARDS net trials, retrospectively analyzed the relationship of barotrauma and various ventilator parameters.[12] Controlling for the covariates, only PEEP and not peak/plateau pressures was associated with an increased risk of barotrauma.

The recommended ventilation strategies in C-ARDS also went through significant controversy based on the varying compliance of C-ARDS (L and H variants) reported in literature (12.13). Our data reflects ventilation practice based on information available at that time in the pandemic. Standard ARDS strategies with moderate PEEP 8 (IQR 6, 10) were used in our cohort, which by itself is less likely to cause barotrauma. The median P-peak and P-plateau were 33.5 cm H2O (IQR: 30, 35.25) and 30 (IQR 25.5, 31) cm H2O, respectively, in our study. Attempts made to keep the Pplat. < 30 cm H2O were challenging in the later phases of disease as the lungs became stiffer and compliance dropped. These pressures are indicative of advanced disease, and present major challenges in optimizing ventilation which may have predisposed to barotrauma.

The explanation for a high incidence of barotrauma in NIV can be partly explained by the longer duration of NIV done for C-ARDS, a strategy not commonly followed in pre-pandemic management of ARDS. The recommended ventilatory protocol was based on experience and expert opinion at that time in the pandemic and varied between ‘early intubation’ versus a ‘delayed intubation’ approach. We followed the ‘delayed intubation’ protocol of maximally extending NIV with careful monitoring prior to iMV, which can also explain the higher incidence of barotrauma on NIV. Some patients also opted for a do-not-ventilate strategy which led to prolonged NIV(N=7). The barotrauma with iMV is easier to explain, as iMV initiation usually happened when the patients were in a progressive or advanced stage of disease.

The barotrauma in our study occurred at a median of 6.5 days (IQR 4.75,13) from admission and 15 days (IQR 10.25,18.0) from symptom onset. The relationship between duration of illness and barotrauma is shown in the graph **(Fig 2)**, showing a higher incidence in the 2^nd^ to 3^rd^ week of illness, suggesting that some patients may have developed an early fibrotic component that might have enhanced the barotrauma risk. This ‘time from disease-onset’ appears to be an important risk factor for barotrauma.

Other probable extrinsic factors for barotrauma include consequences of C-ARDS specific medications, not commonly used in earlier viral pneumonia /prior pandemic therapy. These include medium to high dose steroids uniformly used in all patients as recommended by the Solidarity trial [13] which can enhance pulmonary tissue friability and promote resistant organism superinfection. Both these effects can be expected to augment the primary lung injury of COVID-19.[14] Thus, steroids can not only predispose to barotrauma, but also impair healing when the C-ARDS afflicted lungs are on long-term PPRS/PEEP. The use of immune suppressants such as tocilizumab can also add to impairment of healing and superinfection leading to both barotrauma and delayed healing after the barotrauma.

Intrinsic factors include severity of the ARDS, and related pathology arising from pulmonary vascular aspects. In several studies, the severity of the underlying ARDS was found to be a better predictor of barotrauma than the airway pressures. [6] The median P/F ratio of patients on MV at the time of barotrauma was 116.4 (IQR 72.4, 193.25). A significant number (31.6 %) of patients had P/F ratios below 100, the threshold for defining very severe ARDS. Thus, the severity of the underlying ARDS may have contributed to the increased risk of barotrauma along with the airway pressures in this study. Lemmers et al described a barotrauma incidence of 23 out of 169 C-ARDS patients compared with 3 out of 169 regular-ARDS. The median P/F ratio in the 23 C-ARDS with barotrauma was 105 (IQR 81, 137) compared with 114 (IQR 86, 153) in regular ARDS.[7]

In terms of pulmonary vascular involvement, COVID-19 ARDS is associated with pulmonary vascular micro/macro-thrombi formation, with an increased risk of pulmonary thromboembolism and pulmonary infarction. [15] These thrombotic events have been postulated to cause necrosis and damage to the alveolar membrane and can contribute to subsequent barotrauma. D-dimer is one of the soluble fibrin degradation products that result from breakdown of thrombi by the fibrinolytic system, elevation of which indicates ongoing thrombosis. The median D-dimer levels in the MV patients was elevated at 1381.5 (IQR 742.25, 2285.50) (Normal range < 500 ng/mL) in our cohort, and 3/38 (%) patients had DVT on Doppler ultrasound imaging. These findings are consistent with prior reports of similar effects noted with other coronavirus infections, including SARS-CoV1 and MERS-CoV ARDS.***[7-9]*** Finally, this aspect of elevated D-dimer with thrombosis also complicated management as ICD tubes had to inserted in the presence of therapeutic anticoagulation with an enhanced bleeding risk.

In this study, 24/38 (63.2%) patients with barotrauma including 15/18 (83.3%) on MV with barotrauma succumbed to their illness despite appropriate measures for the barotrauma. The clinical course took a sharp downhill course after the onset of barotrauma with death occurring in a median of 4 days, implying that the onset of barotrauma was a marker for worse prognosis. Barotrauma appears to be an independent risk factor for increased mortality and hospital length of stay in C-ARDS patients. McGuinness G et al. reported a odds ratio of 2.2 for mortality in a cohort of 601 patients with C-ARDS.[5] Especially in the iMV group, interventions such as ICD placement, reduction of PEEP and positive pressure ventilation and steroid reduction had a limited impact, reflecting the advanced nature of the lung damage.

**(L to R) Fig 3.**
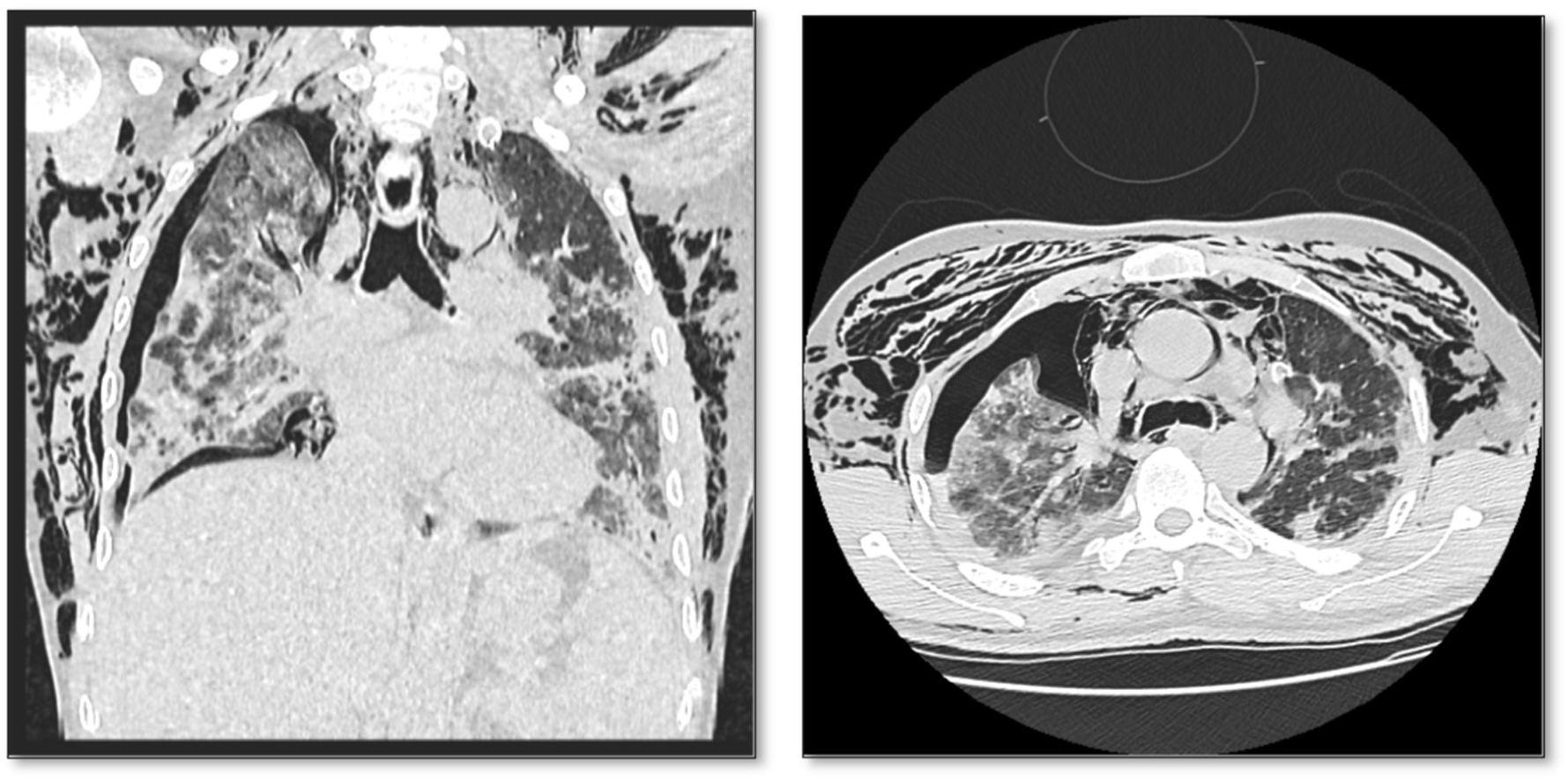
(a). Coronal CT section showing right sided pneumothorax, pneumomediastinum and bilateral extensive subcutaneous emphysema in patient with C-ARDS. Fig 3. (b) Axial CT section with pneumothorax, pneumomediastinum and subcutaneous emphysema

Our study has several limitations. The data in our study is a reflection of a particular strategy, namely initial NIV resorting to iMV as a last resort and reflects the incidence of barotrauma and outcomes with such an approach. In addition to being an expert recommendation, this also was a pragmatic approach considering imminent ventilator and staffing shortages, as iMV and proning requires more intensive resources. Another limitation inherent in this approach is the varying inflation pressures and fluctuating PEEP with NIV, challenges in implementing desired tidal volume and difficulties in precise monitoring. NIV also comes with potential abrupt transitions from ventilation to supportive oxygen and vice-versa as patients are given breaks for compliance, eating and other activities, all of which could be possible risk factors for barotrauma. In addition, as it was not possible to do CT scans in these sick C-ARDS patients, we were unable to define pre-existing or newly developed pulmonary lesions such as emphysema, cysts, underlying fibrotic lung disease, onset of new COVID-related fibrosis and small cavities. The mean Pplat. in our study is the upper limit of normal and reflects the stiff lungs in severe C-ARDS, with likely fibrosis in the patients who were treated beyond 2-3 weeks.

## Conclusion

Our study shows a significant incidence of barotrauma in patients with C-ARDS on positive pressure respiratory support with a rapid downhill course after the occurrence of barotrauma despite intervention, especially in the iMV group. Causes for the barotrauma include both disease-related and treatment related aspects. Though some of these are common to other viral pneumonias, such as MERS and SARS-COV1, there were unique treatment aspects in this pandemic, such as steroids and NIV for ARDS. Measures for prevention and early recognition of barotrauma are important aspects impacting on outcomes in these critically ill patients.

## Data Availability

The data is available on request.

## Acknowledgement

The authors would like to acknowledge the guidance received from Dr. Michael Cutaia MD in the preparation of the manuscript.

